# Development and Usability of a Mobile Ecological Momentary Assessment Platform for Dietary Surveillance in the U.S.

**DOI:** 10.64898/2025.12.10.25341980

**Authors:** Susan M. Schembre, Michelle R Jospe, Rick Weiss, Christopher A. Taylor, Edward J. Bedrick, Jessie Somerville, Julia Felrice, Kelli M. Richardson, Genevieve F. Dunton, Cynthia A. Thomson

## Abstract

Modernizing dietary surveillance is essential for addressing diet-related chronic disease, yet traditional self-report tools remain burdensome and error-prone. *Edna*, a mobile ecological momentary diet assessment (mEMDA) platform, was developed through a multidisciplinary, user-centered process to enable real-time, low-burden dietary reporting with image-assisted portion estimation and automated nutrient coding. In three iterative rounds of national remote testing (N = 146 U.S. adults, 19-65 years), participants were randomized to event- or interval-contingent sampling for 14 days. System Usability Scale (SUS) scores rose from 71.7 to 78.8, surpassing the average benchmark for digital health apps (SUS = 68). Engagement ranged from 92.5-97.6% of study days between rounds, and compliance among interval-contingent users varied between rounds (76-87%). Participants rated portion-size images and navigation as intuitive and culturally inclusive. *Edna* achieved above-average usability and strong engagement, demonstrating the feasibility of scalable, EMA-based dietary surveillance for digital public-health nutrition monitoring.

## Introduction

Dietary surveillance is critical to U.S. public-health efforts because diet-related chronic diseases, including obesity, type 2 diabetes, cardiovascular disease, and several cancers, remain leading causes of preventable morbidity and mortality.^1,2^ Effective surveillance enables timely and accurate data at the population-level. National dietary data serve as exposure measures for etiologic research, calibrate measurement error, enable dose-response and risk assessments, validate dietary biomarkers, and support impact evaluations and policy counterfactuals.^3,4^ Although advanced dietary assessment methods such as image-based approaches, biomechanical sensors, and metabolomics offer promise, they remain inaccessible and insufficiently validated for use in population-level surveillance.^5–8^ Accordingly, public health surveillance will depend on modernized self-report tools that can produce high-quality, population-scale dietary data with the rigor necessary to drive effective action against diet-related disease.

Traditional dietary surveillance tools such as 24-hour dietary recalls (**24HR**) and food frequency questionnaires (**FFQ**), which ask people to report foods consumed over the past 24-hours (24HR) to up to one year (FFQ), are widely used due to their practicality and scalability.

However, they suffer from several well-documented limitations, including recall bias, social desirability bias, respondent burden, and inaccuracies in portion size estimation, particularly when used with diverse or low-literacy populations.^9–11^ Ecological momentary assessment (**EMA**) offers a strategy whereby individuals complete brief, repeated surveys in real or near real-time using mobile devices.^12^ When applied to diet, EMA allows logging of intake soon after consumption, thereby shortening recall intervals, reducing memory load, and minimizing reactivity. Mobile ecological momentary diet assessment (**mEMDA**) can also capture important contextual factors, such as location, social setting, or emotional state, support cross-cultural comparisons through flexible food lists, and streamline researcher workload via automation.^13^ Despite these advantages, mEMDA tools and sampling strategies have not yet been systematically optimized or validated for population-level dietary surveillance, and even fewer have been evaluated for usability and engagement as surveillance tools; key determinants of sustained compliance and data quality.^13,14^

Recent advances in mobile health technology make near real-time, low-cost dietary data collection feasible at scale. In contrast, traditional surveillance methods remain costly, periodic, and poorly equipped to capture the dynamic nature of dietary intake throughout the day.^15^ We hypothesize that mEMDA can offer a practical path to continuous monitoring by delivering timely prompts, guiding users through intuitive, self-administered logging interfaces, and streaming structured data directly into nutrient databases for rapid analysis. Supporting evidence for the feasibility, acceptability, and potential utility of mEMDA in diverse populations is accumulating.

Our 2018 systematic review of 20 mEMDA protocols reported mean prompt response rates of 79% and day-level concordance with 24HR exceeding 80%.^13^ A 2022 review of 39 studies in adolescents and young adults (16–30 years) focusing on eating behaviors, dietary intake, and related contextual factors found compliance rates of at least 80% in most studies and generally favorable usability ratings.^16^ Most recently, a 2024 pooled analysis of four intensive EMA studies in older adults (mean age ≈72 years) that tracked eating occasions and dietary patterns documented an average prompt compliance rate of 77.5%, demonstrating feasibility even among populations often considered less tech-savvy.^17^

Building on this evidence base, we developed ***Edna***, a research quality, EMA-based Diet and Nutrition Assessment tool. *Edna* is a mEMDA app for use on smartphones that was engineered for large-scale, low-burden dietary surveillance across the adult life course. *Edna* integrates real-time self-report with an image-supported, culturally inclusive food list and automated linkage to a publicly available nutrient database. Key design goals included minimizing user fatigue, reducing time burden, shortening recall intervals to lessen reliance on memory, ensuring broad accessibility, and leveraging visual cues to improve portion-size estimation. In this paper we describe *Edna’s* user-centered and data-driven development and an iterative usability study designed to finalize the platform’s interface and feature set. As the first report in a planned series on *Edna’s* strengths and limitations, this paper evaluates *Edna’s* usability and acceptability in a socio-demographically and regionally diverse sample of U.S. adults. Findings will guide subsequent validation studies and inform future design refinements to support integration of mEMDA into routine public health nutrition surveillance.

## Methods

### Conceptual Rationale for *Edna*

*Edna* was purpose-built to narrow the gap between traditional self-report dietary assessment tools and emerging approaches that are more objective, while remaining practical for large-scale surveillance. Developed through an iterative, user-centered design process, the platform was shaped by a multidisciplinary team including behavioral scientists, registered dietitians, software engineers, and graphic designers. Its architecture was informed by three sources of formative evidence: (1) findings from our systematic review of mobile EMA methods; (2) analyses of nationally representative dietary intake data used to guide the development of a culturally and regionally diverse food list; and (3) rapid-cycle prototype testing with prospective end users. Together, these activities informed the creation of a smartphone-based tool that combines a demographically inclusive food list with streamlined meal-entry and image-based portion estimation pathways, and automated nutrient coding through a publicly available database. The current version of *Edna* represents a proof-of-concept platform focused on a targeted set of nutrients of concern (saturated fat and added sugars), with plans to expand its nutrient coverage in future iterations.

The design of *Edna* was guided by the overarching goal of improving the feasibility and accuracy of dietary surveillance methods for population-level research. To achieve this, we pursued two primary strategies. First, we incorporated EMA to reduce the recall interval and lessen dependence on memory compared to traditional dietary assessment methods, while also minimizing the need for self-initiated reporting by leveraging system-triggered prompts. Second, we prioritized minimizing participant burden, recognizing that shorter and cognitively simpler interactions can improve compliance and enhance data quality.

EMA was selected over conventional recall methods to enable real-time or near real-time capture of dietary intake, which enhances ecological validity and supports longitudinal tracking of usual intake alongside other time-varying behaviors. It facilitates the simultaneous collection of contextual variables (e.g., social setting, mood, location) that can help illuminate the antecedents and consequences of dietary choices. Although reactivity to self-monitoring can occur when individuals consciously attempt to change the measured behavior or complete reports prospectively, retrospective recording (e.g., documenting food intake after meals) shows little to no evidence of behavioral change from repeated EMA prompts.^12,18^

*Edna* was specifically designed to address persistent limitations of traditional methods such as 24HR and FFQ. These tools are time-intensive (often requiring 25 to 60 minutes per assessment), cognitively demanding, and prone to memory bias. They are also resource-intensive for researchers (e.g., 24HR) or may be limited in their ability to accommodate cultural food diversity or support meaningful comparisons across populations (e.g., FFQ). In contrast, *Edna* was built to require less than 15 minutes of participant time per day, reduce memory and cognitive burden, and minimize administrative overhead through fully automated data capture and nutrient coding.

Accessibility and inclusivity were foundational to *Edna’s* development. The food and beverage list was derived from the National Health and Nutrition Examination Survey (**NHANES**), a nationally representative surveillance system of dietary and health-related behaviors in the U.S. population to ensure coverage of items commonly consumed by U.S. demographic subgroups.

Visual simplicity and clarity were prioritized throughout the interface, and a color-blindness simulator was used during design testing to ensure accessibility for users with color vision deficiencies.

### Technical Specifications

*Edna* was developed in partnership with Viocare, Inc., a National Institutes of Health (**NIH**)-funded diet-assessment software company, through an iterative, user-centered process that paired behavioral scientists and dietitians with graphic designers and software engineers. The mobile app was built using the React Native Expo framework, which enables cross-platform development from a single codebase while maintaining the look and feel of native applications. Platform-specific binaries are packaged for iOS and Android and distributed through the Apple App Store and Google Play, meeting each platform’s privacy, security, and accessibility standards.

The backend is built on Firebase, a cloud-based infrastructure that supports secure authentication, data storage, and backend processing. Google Sign-In provides user authentication, and Firestore, a scalable NoSQL database, stores participant data in real time. Cloud Functions are used to handle background operations such as notification scheduling and data processing, while Firebase Storage manages user-generated content with granular access controls. The system is fully managed, scalable, and secured using Firebase’s authentication and security rules.

*Edna* supports both interval- and event-contingent EMA sampling strategies, allowing researchers to tailor data collection schedules to study needs. Logged dietary entries are stored using unique *Edna* food codes, which are mapped to NHANES-based food items and recipes through the United States Department of Agriculture (**USDA**) Food and Nutrient Database for Dietary Studies (**FNDDS**). This mapping allows for the generation of nutrient output files with time-stamped entries, enabling analysis by eating occasion and day.

A secure, web-based administrative dashboard provides investigators with role-based access to manage participants, monitor engagement, review system logs, and export nutrient data. The dashboard, built with React JS, is integrated with the same backend services that support the mobile app, allowing for coordinated updates and streamlined data management. The system architecture was designed to accommodate future expansion, including the addition of new nutrient modules and sampling schedules, with minimal redevelopment effort.

### Food List

*Edna’s* food list was developed using a structured, data-driven process aimed at improving the precision dietary assessment in U.S. adults. As detailed in our recent analysis of NHANES 2005–2018 data, we identified the leading food and beverage contributors to the nutrients targeted at this stage of development (saturated fat and added sugar intake) by aggregating two days of 24-hour dietary recall data from over 36,000 adults. Each reported item was mapped to USDA “What We Eat in America” third-level food categories and ranked by its proportional contribution to the targeted nutrient intakes, both in the general population and across key sociodemographic subgroups. Food categories were retained if they contributed to the top 90% of daily saturated fat and added sugar intakes at the population or subgroup level. This process resulted in 95 unique food categories that accounted for ≥90% of saturated fat and added sugar consumption for more than 88% of the adult U.S. population.17 Despite the narrow list of targeted nutrient, the resulting list of foods accounted for ≥91% of total protein intake, ≥82% of total carbohydrate intake, and ≥95% of total dietary fat intakes across subgroups of the adult U.S. population supporting the future expansion of *Edna* as a comprehensive dietary surveillance tool.

To prioritize foods most relevant to daily intake estimation, we applied a 2-gram inclusion threshold by retaining foods that typically contribute at least 2 grams of saturated fat or added sugars per standard portion. This cutoff was derived from dietary guidelines recommending no more than 10% of daily energy from saturated fat (∼20 grams/day on a 2,000 kcal diet), allowing us to focus on items with a meaningful nutritional impact. In cases where a nutrient-dense food (e.g., cheese) was commonly consumed with a low-nutrient “vehicle” (e.g., crackers), we included both to support accurate meal reconstruction, even when the vehicle food alone fell below the threshold. This pragmatic strategy helped preserve ecological validity while maintaining the overall parsimony of the list.

The 95 food and beverage categories were organized into a hierarchical structure comprising 12 broad food groups (Level 1), approximately 350 narrower food groups (Level 2), and over 1,000 unique food types (Level 3).^19^ Decisions to include specific foods and preparation styles were data-driven, based on expert review by a team with expertise in nutrition and dietary assessment methods and the estimated amounts of saturated fat and added sugars contributed by standard portion sizes, using nutrient values from the USDA FNDDS database. This structure enabled intuitive navigation while allowing for nuanced customization. All entries were ultimately linked to FNDDS nutrient profiles through a structured recipe database, enabling real-time nutrient coding and minimizing post-processing burden.

### User Interface and User Experience Design (UI/UX)

#### Overview

*Edna’s* user interface was developed to reflect established practices in EMA while prioritizing ease of use and scalability for population-based research. Prompts were included as a standard feature of interval-contingent EMA protocols, and reminders were incorporated in the event-contingent design based on evidence that they support compliance. Image-assisted portion size selection, based on Viocare’s Vioscreen FFQ,^20^ was used to improve estimation accuracy, and additional features, such as guided food search and a favorites list, were designed to reduce burden and streamline the reporting process. The sections that follow describe the structure and functionality of *Edna’s* user interface, including the entry experience by sampling condition, food and portion size selection, features that support completeness and engagement, and the onboarding and in-app support resources available to users. Screenshots of the final app can be seen in **Figure 1**.

**Figure 1.**
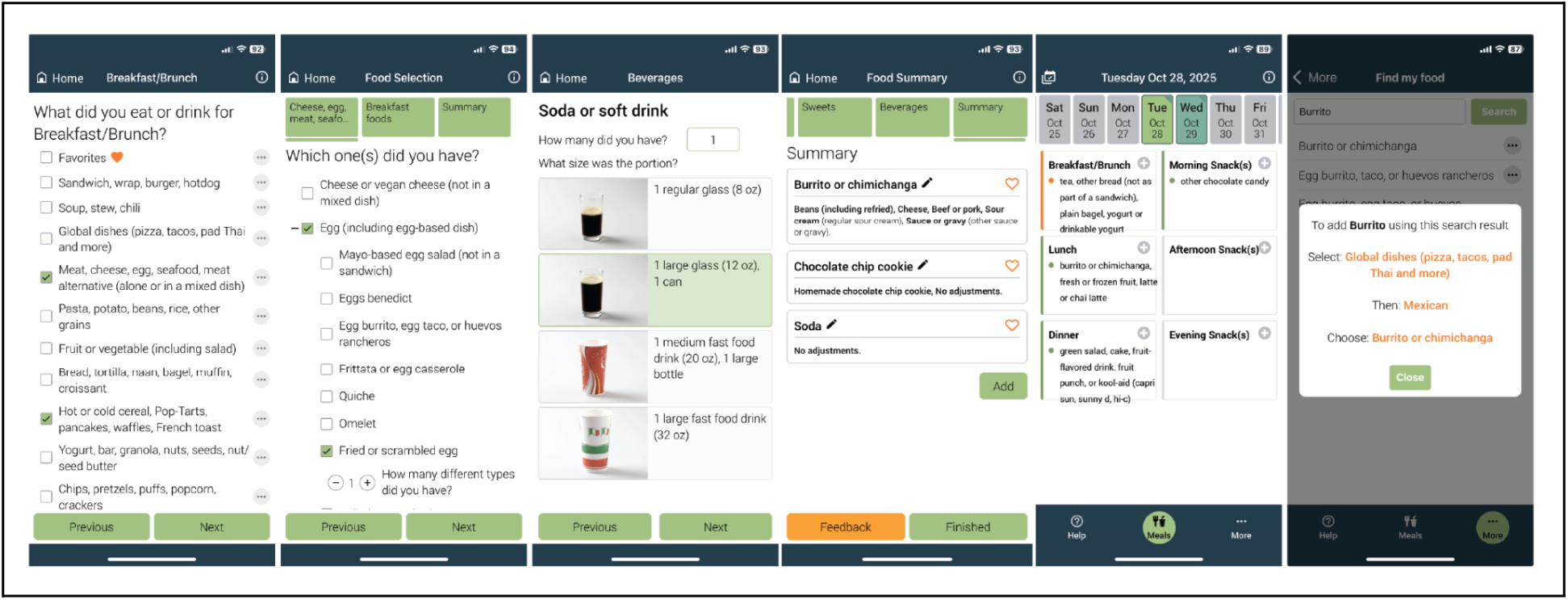
Screenshots of the *Edna* app

#### Initiating Food Entry by Sampling Condition

*Edna’s* interface accommodates event-contingent and interval-contingent sampling. While the core logging workflow is consistent across groups, the entry experience differs at the outset. Event-contingent users are instructed to self-initiate logging after each eating episode. Their home screen displays a grid of clickable tiles labeled by meal type (breakfast/brunch, lunch, dinner, morning snack, afternoon snack, evening snack). In contrast, interval-contingent users receive automated in-app notifications at pre-specified intervals throughout their specified waking hours. The first prompt of the day is delivered at the self-reported wake time, and subsequent prompts are sent every 2.5 hours throughout the day, with the final prompt occurring 30 minutes before the self-reported bedtime. Each prompt asks whether the participant consumed any food or beverage during the previous 2.5-hour interval. For example, a prompt might ask, “Did you eat or drink anything between 9:30 AM and 12:00 PM?” If the user selects “no,” the app records that no eating occurred and no further action is required. If the user selects “yes,” they are guided into the food entry workflow. If no response is made within 10 minutes of the prompt, the app delivers up to two follow-up reminders to encourage timely reporting. Rather than meal labels, the interval contingent home screen presents a vertical list of labeled time windows that correspond to the interval of time preceding the prompt. Both workflows prompt users to enter the approximate time of the eating event before proceeding to food and beverage entry. All users can access a scrolling calendar at the top of the home screen to edit or add entries for the current day or the previous day and to review past days. These structured approaches support near real-time data capture while accommodating individual daily schedules.

#### Food Entry and Portion Size Selection

*Edna’s* food logging interface is designed to support accurate and efficient self-reporting through a structured, low-burden workflow (Figure 1). After selecting a meal label or time interval and entering a meal time, users begin food entry by choosing from 12 broad food categories (Level 1). Each selection dynamically reveals a smaller set of relevant subcategories (Level 2), which in turn lead to a curated list of specific food types (Level 3). This nested structure ensures that users are only shown contextually relevant options, streamlining the process and minimizing cognitive load. Once a food type is selected, users are prompted to specify preparation details (for example, protein or sauce ingredients) and any additions or condiments included after preparation. For example, a Thai chicken curry dish topped with a fried egg would be logged as: Level 1: Global dish; Level 2: Asian or Indian; Level 3: Curry or masala dish, with chicken and a coconut-based curry sauce, and an added egg.

All primary foods and beverages require portion size selection. Certain condiments and additions also require portion selection, particularly when they have variable serving sizes that could meaningfully impact saturated fat or added sugar intake (such as whipped cream, frosting, or gravy). Portion size is estimated using labeled photographs tailored to the selected item (image-assisted portion size estimation). For foods with a standard portion size (for example, eggs or granola bars), a single portion image is shown and clearly labeled (such as “1 whole or scrambled egg”). Users are asked to report how many servings of that portion they consumed. For foods without a standard serving size (such as casseroles, ice cream, or pasta), multiple portion size images are displayed in a vertically scrollable format. Each image is labeled for clarity (such as “1 scoop or ½ cup” or “1 cup”), and users select the image that best reflects the amount consumed. This flexible, image-supported approach is meant to improve estimation accuracy and consistency across diverse food types.

Completed entries are displayed on a summary screen that presents a plain-language overview of the selected food or beverage, including preparation details and portion size. From this screen, users can make real-time edits, add a forgotten food or beverage to the current entry, or mark the item as a favorite for quicker access in the future. This review step supports accuracy and completeness while giving users control and flexibility before finalizing each submission.

#### Features Supporting Completeness, Compliance, and Feedback

To reduce missing data and support consistent engagement, *Edna* includes several features designed to promote adherence, reduce user burden, and enhance the accuracy of logged entries.

*Prompts and reminders*. Interval-contingent users receive automated in-app notifications based on their reported sleep and wake times. Users can enter different weekday and weekend schedules to reflect their typical routines. Event-contingent users are instructed to self-initiate logging after each eating occasion. To support regular engagement and reduce reliance on memory, *Edna* allows users in this group to set custom reminders aligned with their typical meal and snack times. These reminders can be enabled, disabled, or modified at any point. This feature preserves the flexibility of event-contingent logging while offering gentle prompts to encourage consistent use.

*Favorite foods*. To streamline data entry and support repeated reporting of common items, *Edna* includes a favorite foods feature. Users can mark frequently consumed or leftover items as favorites, which are then stored in a personalized list for quick access in future logging sessions. Favorite foods retain all previously specified details, including preparation method and portion size, but can be edited as needed before submission. This feature reduces repetitive entry, improves user efficiency, and supports consistency across repeated meals and snacks.

*Navigation assistance*. To support accurate food identification and reduce navigation errors, *Edna* offers a built-in “Find My Food” feature. This tool provides step-by-step instructions to help users locate items by name, food group, or common alias. Alias terms were generated using generative AI to account for regional, cultural, and colloquial naming variations. For example, searching for “pop” directs users to soft drinks, and “Pad Thai” links to the broader category of Asian noodle dishes. By improving searchability and guiding users toward the correct entry path, this feature reduces user frustration and helps maintain completeness and consistency across records.

Together, these features are designed to enhance usability and data quality across both sampling conditions. They work in concert to support a more seamless, participant-centered logging experience while minimizing burden and encouraging sustained engagement over time.

#### Onboarding and In-App Support

To ensure users are able to navigate the app and complete entries with confidence, *Edna* includes a suite of onboarding and in-app support resources. Upon first login, all users are guided through a series of onboarding screens that introduce the app’s layout, navigation flow, logging procedures, and prompt response expectations. These onboarding materials were designed to set clear expectations and reduce early user confusion, particularly among those unfamiliar with dietary tracking tools. The onboarding tutorial remains accessible via a Help menu, allowing users to revisit the content as needed.

Additional support features are embedded within the app to assist users in real time. These include video demonstrations of the food logging process, a searchable FAQ section, and a Contact Us feature that allows users to reach the study team or app developers for assistance. These tools were developed to address both common and emergent user needs, from clarifying app functions to resolving technical issues.

Together, these onboarding and support features are intended to promote confidence, reduce technical barriers, and minimize attrition due to confusion or frustration. By combining structured orientation with on-demand support, *Edna* aims to create a user experience that is both intuitive and responsive.

### Usability Testing and Iterative UI/UX Refinement

#### Study Design and Setting

This study used a sequential, multimethod design to iteratively evaluate and refine *Edna*. Usability testing was conducted remotely in the U.S. between June 2024 and June 2025, with each round involving 14 days of app use per participant. The study design was intentionally iterative, allowing the research team to modify the app between rounds based on user feedback and performance metrics. Usability testing was conducted in iterative rounds until usability scores plateaued, defined as a change of < 3 points between rounds, with a maximum of three rounds. The study was approved by the Georgetown University Institutional Review Board.

#### Study Participants

Participants were adults aged 19 to 65 years residing in the U.S., recruited remotely through Research Match, a volunteer registry supported by the U.S. National Institutes of Health as part of the Clinical Translational Science Award program (https://www.researchmatch.org/researchers/faq). Eligible participants were required to own a smartphone capable of downloading and running the *Edna* app and be able to read and understand English. Individuals were excluded if they reported a current or past diagnosis of an eating disorder or a BMI below 18.5 kg/m², based on self-reported height and weight. Although the study did not target vulnerable populations, individuals from a range of socioeconomic and demographic backgrounds were eligible to participate.

To ensure demographic diversity and broad applicability, the study employed a stratified recruitment strategy across age, sex, BMI category, and geographic region. Prospective participants were screened for eligibility and enrolled using REDCap, a secure, web-based platform. Informed consent was obtained electronically prior to participation.

#### Procedures

##### *Edna* Food Entry Workflow Usability

Following electronic informed consent, participants were enrolled remotely and randomly assigned to one of two sampling conditions: event-contingent or interval-contingent. Participants were instructed to download the *Edna* app from the Apple App Store or Google Play Store and were provided with a study-specific username and a unique password to register the app. Once registered, participants could begin logging food and beverage intake using the flow corresponding to their assigned sampling condition. Study staff monitored the administrative dashboard to confirm successful registration and followed up with participants who had not registered within the first 48 hours. Participants who failed to register were contacted up to three times with reminders and trouble shooting queries. Only those who registered the app were able to participate in study activities.

Participants were asked to log all meals, snacks, and beverages consumed over a 14-day period. A 14-day use period was selected to assess short-term engagement under naturalistic conditions and to explore whether participants would continue using *Edna* beyond the typical 5-to 7-day duration commonly used for consecutive food recording in dietary studies. No additional training or support was provided beyond the onboarding and help features embedded within the app, allowing for a realistic assessment of standalone usability.

Technical issues were reported by participants through the in-app feedback form, daily REDCap surveys, or direct, email communication with the study team including Viocare personnel. Study staff monitored participant activity daily during the 14-day period using the *Edna* administrative dashboard. App bugs, crashes, and trouble shooting requests were addressed in real time, with any necessary updates deployed without requiring reinstallation. This allowed participants to continue logging without disruption while enabling ongoing refinement of the app between testing rounds.

Structured usability feedback was also collected at three time points using custom REDCap surveys. On the day of registration, participants completed an onboarding feedback form that prompted open-ended reflections on their experience downloading and setting up the app under multiple feedback categories (“Downloading the app,” “Tutorials/Videos,” “Bug reports”) and submitted additional comments as option, open text. During the 14-day use period, participants received daily feedback forms via text, which asked them to categorize their feedback (e.g., “Missing food,” “Suggestions for improvement,” “Bug report”) and provide open-ended comments about their experience that day. These reports offered real-time insight into usability challenges and supported early identification of friction points in the logging workflow.

On day 15, participants were asked to complete end-of-study feedback surveys, including the System Usability Scale (**SUS**). This included a comprehension check about *Edna’s* intended purpose, followed by Likert-style items on the usefulness of the onboarding materials and support features. Open-ended questions captured what participants liked and disliked, identified foods that were difficult to enter, and gathered suggestions for improving the search and entry experience. Users were reminded to complete each of the end of study tasks at least three times before they were considered lost-to-follow-up. The same 14-day protocol was used for all rounds. After each round, participant feedback and survey data were reviewed to guide iterative app refinements.

##### Portion Size Estimation Usability and Food Selection Accuracy (Final Round Only)

As part of the final round of testing, a subsample of participants completed a simulated food logging task designed to assess the accuracy and usability of *Edna’s* portion size estimation workflow. Each participant was provided a single, one day menu comprising three meals and one snack. Menus included photographs of each food or beverage in standardized portions. Participants were asked to enter all menu items into the *Edna* app in a single sitting while in a Zoom session with a trained study team member, selecting both the correct food and the most appropriate portion size image. The goal was to assess whether users could successfully navigate the logging interface and accurately match foods and portion sizes.

Participants were prompted by study staff to determine what their usual portion size for each food item would be before choosing the portion size image and how many they would have and entering it into the mobile tool. Study staff recorded the portion size image selection and whether the selection matched their usual portion size for this food. Study staff also noted whether the user experienced any technical difficulties with entering portion sizes on the mobile tool.

Following the simulation, participants completed a structured questionnaire to evaluate the usability of *Edna’s* portion size estimation interface. This survey included 5-point Likert-scale items assessing clarity and relevance of portion size images, ease of use, alignment with typical eating behaviors, and overall satisfaction. Additional open-ended items captured specific suggestions for improving the portion size flow, including terminology, labeling, and image presentation. The final item asked participants to rate how satisfied they would be if their suggested changes were implemented.

##### Participant Compensation

Participants were compensated for their time and engagement, with compensation schemes varying slightly by study round. In Round 1, participants could earn up to $70, calculated as $5 per day of *Edna* use over the 14-day period, multiplied by their percentage of engagement (i.e., the proportion of days they logged at least one food or beverage entry). In subsequent rounds, compensation increased to a maximum of $80, including $1 for registering the app, $1 per day (up to 14 days) for logging foods and beverages consumed, $30 for completing the end-of-study feedback form, and $35 for completing the SUS. Participants who were randomized to complete the portion size testing task did not receive additional compensation. All payments were issued electronically after participation was verified.

#### Measures

##### Usability

The SUS was the primary quantitative usability outcome, and a predefined stopping rule was used to determine whether additional testing rounds were needed. The SUS, a validated 10-item questionnaire that yields a composite score from 0 to 100 on the general usability of a wide range of systems, including software, with higher scores indicating better perceived usability.^21^ We planned for a minimum of two rounds of usability testing and app refinement with a goal of achieving an average SUS score of 80.8. Specifically, further rounds would be conducted only if SUS scores fell below a mean threshold of 80.8 in either sampling condition. This goal corresponds to a usability rating of A and above on the commonly used Sauro–Lewis SUS curved grading scale, indicating high user satisfaction and strong product usability.^22^ Additionally, open-ended responses from an onboarding feedback survey, daily feedback surveys, end-of-study feedback form, and the portion size usability questionnaire were analyzed to identify usability barriers, navigation issues, and opportunities for improvement. Surveys were administered on day 15 via REDCap.

##### Engagement

Participant engagement was assessed using backend data from the *Edna* administrative dashboard. Engagement was defined as the percentage of the 14 study days on which a participant logged at least one food or beverage entry. This metric was calculated for all participants and used to characterize usage patterns and frequency of app interaction. Engagement was compared between the interval-contingent and event-contingent groups to examine whether prompting style influenced sustained use.

##### Compliance

Compliance was assessed for participants in the interval-contingent group only, who received automated prompts at scheduled intervals throughout the day. Compliance was defined as the percentage of intervals completed, meaning intervals in which the participant either reported not eating or submitted a food or beverage entry. This metric captured adherence to the structured EMA protocol over the 14-day period.

##### User Satisfaction

Additional feedback on the satisfaction and usefulness of specific app features, including the onboarding tutorial, video tutorials, and the portion size selection process was obtained with 5-point Likert scales where higher scores reflected more favorable responses.

### Statistical Analysis

#### Randomization and Sample Size Justification

Participants were recruited using stratified sampling to ensure demographic diversity across key characteristics, including self-reported gender, age group (19–35, 36–50, 51–65), BMI category (18.5–24.9, 25–29.9, ≥30), and U.S. geographic region (Northeast, Southeast, Midwest, Southwest, West). Stratification was used to ensure broad geographic and demographic representation, with a minimum of five participants targeted from each region per round.

Participants were randomly assigned at enrollment to one of two sampling conditions (event-contingent or interval-contingent) using block randomization (block sizes of 2, 4, or 6) created with https://www.sealedenvelope.com/simple-randomiser/v1/lists and implemented through REDCap. In the final round of testing, a subsample of participants volunteered to also be randomized to one of three standardized simulated menus used in the portion size estimation task.

The study aimed to enroll approximately 50 participants per round, for a total sample size of up to 150 participants across three rounds of testing. This sample size was selected to balance feasibility with analytic precision and was sufficient to achieve thematic saturation for qualitative feedback within each testing wave. It also supported planned exploratory analyses comparing SUS scores by sampling condition and across study rounds. With approximately 75 participants per condition, the study had 80% power to detect a moderate effect size (Cohen’s d ≈ 0.5) in SUS score comparisons between groups using two-sided tests at α = 0.05. If no differences in SUS scores were observed between sampling conditions, scores were pooled to examine changes in usability across rounds. The staged design allowed for iterative refinement of the *Edna* interface while preserving consistency in core study procedures. We fit two linear regression models to examine demographic and study-related predictors of usability and engagement outcomes. The first model included SUS score as the dependent variable, and the second model included percentage engagement. Both models specified survey round, randomization protocol, sex, education, age, race, ethnicity, BMI category, and U.S. region as predictors of SUS scores.

#### Quantitative Data Analysis

Analyses were stratified by study round and sampling condition. Quantitative data were included only for participants who used *Edna* for at least 3 days during the 14-day testing period. Descriptive statistics were reported as means and standard deviations. SUS scores and app engagement were compared by sampling condition using independent samples t-tests, and overall differences across rounds were assessed using a one-way ANOVA. Secondary outcomes included compliance (interval-contingent sampling only) and feature-specific feedback metrics. Analyses were conducted using R (v4.2.2), and missing data were handled using complete-case analysis.

#### Qualitative Data Analysis

Open-ended survey responses from the onboarding survey, daily feedback surveys, end-of-study feedback form, and portion size usability questionnaire were analyzed by one study team member using inductive coding. Responses were first reviewed line by line to identify recurring themes and patterns related to usability barriers, navigation issues, and opportunities for improvement, without imposing a predefined framework. Generative AI-assisted coding was then applied as a second pass to confirm consistency of theme assignment across the dataset.

Discrepancies between manual and AI coding were reviewed and resolved through discussion among the research team. Comments were categorized by issue type (e.g., difficulty finding foods, unclear portion size labels, confusion with onboarding flow), and repeated comments were tallied in Excel to identify the most commonly cited concerns. After the data were organized, the Principal Investigator and the Viocare project manager reviewed the summarized feedback and jointly determined a course of action for improving the *Edna* platform. Modification decisions were prioritized to address issues cited most frequently. User interface and workflow refinements were made between rounds during the iterative development phase. This rapid-cycle, user-centered approach allowed the study team to incorporate direct participant input into ongoing refinement while maintaining a consistent 14-day testing protocol across all rounds.

## Results

A total of 150 participants were enrolled in the study, distributed across multiple rounds of testing. Those with fewer than 3 days of app use (n = 2) and/or who were missing SUS scores (n = 4) were excluded, resulting in an analytical sample of N=146. Participants across the three rounds were generally representative of the U.S. population of adults aged 19-65 years with respect to age, Hispanic origin, and regional population estimates based on 2024 U.S. census estimates. Females were slightly overrepresented at 57.5% compared to the national estimate of 50.5% and individuals with obesity were slightly underrepresented at 37.2% vs. 40.3% according to the 2024 Centers of Disease Control estimates. The sample was also representative of the U.S. race distribution with modest variations due to more participants identifying as a race other than the presented, single race options. No significant differences were observed across rounds.

**Table 2** presents SUS scores and engagement by study round and sampling condition. Mean SUS scores increased slightly across rounds, from 71.7 (SD = 23.0) in Round 1 to 78.8 (SD = 19.3) in Round 3. There were no significant differences in SUS scores between rounds (p = 0.187) or between the event- and interval-contingent groups within each round (all p ≥ 0.315).

**Table 1.** Participant characteristics by round (N=146)

|  | All | Round 1 | Round 2 | Round 3 | P-value<sup>1</sup> |
| --- | --- | --- | --- | --- | --- |
| N | 146 | 46 | 45 | 55 |  |
| Age, years (mean (SD)) | 41.0 (12.7) | 42.0 (14.2) | 43.9 (12.6) | 38.0 (11.0) | 0.060 |
| Sex (%) |  |  |  |  | 0.698 |
| Female | 84 (57.5) | 28 (60.9) | 26 (57.8) | 30 (54.5) |  |
| Male | 61 (41.8) | 17 (37.0) | 19 (42.2) | 25 (45.5) |  |
| Other | 1 (0.7) | 1 (2.2) | 0 (0.0) | 0 (0.0) |  |
| Race (%) |  |  |  |  | 0.632 |
| White | 105 (71.9) | 36 (78.3) | 31 (68.9) | 38 (69.1) |  |
| Black/African American | 20 (13.7) | 5 (10.9) | 8 (17.8) | 7 (12.7) |  |
| American Indian/Alaskan Native | 2 (1.4) | 0 (0.0) | 0 (0.0) | 2 (3.6) |  |
| Asian | 1 (0.7) | 0 (0.0) | 1 (2.2) | 0 (0.0) |  |
| Native Hawaiian/Pacific Islander | 1 (0.7) | 0 (0.0) | 1 (2.2) | 0 (0.0) |  |
| Other | 17 (11.6) | 5 (10.9) | 4 (8.9) | 8 (14.5) |  |
| Ethnicity (%) |  |  |  |  | 0.219 |
| Hispanic or Latinx | 32 (21.9) | 6 (13.0) | 10 (22.2) | 16 (29.1) |  |
| Non-Hispanic or Latinx | 113 (77.4) | 39 (84.8) | 35 (77.8) | 39 (70.9) |  |
| Unknown | 1 (0.7) | 1 (2.2) | 0 (0.0) | 0 (0.0) |  |
| BMI category (%) |  |  |  |  | 0.196 |
| 18.5–24.9 kg/m<sup>2</sup> | 44 (30.3) | 14 (31.1) | 14 (31.1) | 16 (29.1) |  |
| 25–29.9 kg/m<sup>2</sup> | 47 (32.4) | 11 (24.4) | 20 (44.4) | 16 (29.1) |  |
| ≥30 kg/m<sup>2</sup> | 54 (37.2) | 20 (44.4) | 11 (24.4) | 23 (41.8) |  |
| Education (%) |  |  |  |  | 0.714 |
| High school diploma, GED, or some college | 21 (14.4) | 6 (13.0) | 6 (13.3) | 9 (16.4) |  |
| Associate's or bachelor's degree | 64 (43.8) | 17 (37.0) | 22 (48.9) | 25 (45.5) |  |
| Graduate degree | 61 (41.8) | 23 (50.0) | 17 (37.8) | 21 (38.2) |  |
| Partnered (%) | 105 (71.9) | 32 (69.6) | 34 (75.6) | 39 (70.9) | 0.799 |
| Geographic region (%) |  |  |  |  | 0.873 |
| Midwest | 27 (18.5) | 8 (17.4) | 10 (22.2) | 9 (16.4) |  |
| Northeast | 28 (19.2) | 9 (19.6) | 11 (24.4) | 8 (14.5) |  |
| Southeast | 45 (30.8) | 15 (32.6) | 10 (22.2) | 20 (36.4) |  |
| Southwest | 17 (11.6) | 5 (10.9) | 6 (13.3) | 6 (10.9) |  |
| West | 29 (19.9) | 9 (19.6) | 8 (17.8) | 12 (21.8) |  |

1. P-values for between-round differences were calculated using ANOVA for continuous variables and Pearson's chi-square test for categorical variables. Fisher's exact test was applied to sex and race due to low cell counts. The underweight category was excluded from BMI analyses.

**Table 2.** System Usability Scale (SUS) Scores by round and sampling condition (N=.

|  | Round 1 |  |  |  | Round 2 |  |  |  | Round 3 |  |  |  |  |
| --- | --- | --- | --- | --- | --- | --- | --- | --- | --- | --- | --- | --- | --- |
|  | All (n=46) | Event (n=24) | Interval (n=22) | within round p-value | All (n=45) | Event (n=23) | Interval (n=22) | within round p-value | All (n=55) | Event (n=25) | Interval (n=30) | within round p-value | Between round p-value |
| **Engagement** (% days out of 14), mean (SD) | 93.0 (17.8) | 89.0 (22.6) | 97.4 (9.2) | 0.103 | 97.6 (7.5) | 97.2 (9.3) | 98.1 (5.0) | 0.705 | 92.5 (15.6) | 95.1 (9.2) | 90.2 (19.2) | 0.223 | 0.168 |
| **SUS Score**, mean (SD) | 71.7 (23.0) | 72.2 (20.8) | 71.2 (25.6) | 0.893 | 77.6 (17.7) | 77.0 (17.9) | 78.3 (17.9) | 0.803 | 78.8 (19.3) | 81.6 (16.4) | 76.4 (21.5) | 0.315 | 0.187 |

Note: SUS scores in Round 1 corresponded to a grade of C+ (range: 71.1–72.5). Scores in Rounds 2 and 3 correspond to a grade of B+ (range: 77.2–78.8).<sup>23</sup> Round 3 event sampling met the predetermined threshold of $\geq 80.8$ .

Engagement was consistently high across all rounds, with mean engagement ranging from 92.5% (SD = 15.6%) in Round 3 to 97.6% (SD = 7.5%) in Round 2. Differences by round (p = 0.168) and sampling condition (all p ≥ 0.103) were not statistically significant. When we examine engagement in rounds 2 and 3, which had the same compensation scheme, engagement was consistently high, with those in the event-contingent sampling condition having slightly greater engagement (difference of 2.6 percentage points) which was not statistically significant (p = 0.306) (**Figure 2**). Compliance with interval-contingent prompts was assessed only for participants in the interval group. Data were unavailable in Round 1. In Round 2, the mean compliance was 87.4% (SD = 15.0%), while in Round 3, it was 76.0% (SD = 24.5%).

**Figure 2.**
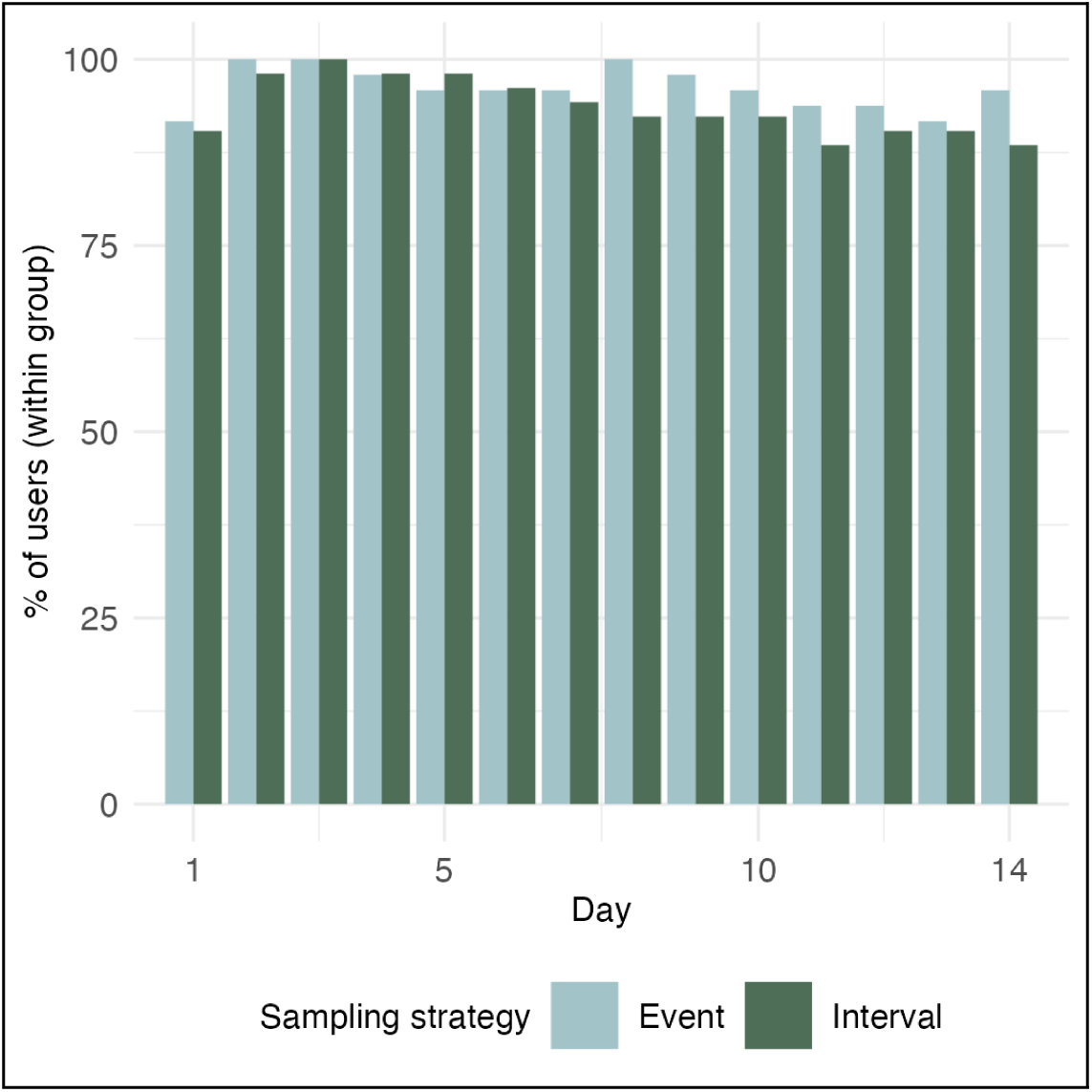
Daily engagement by sampling condition in rounds 2 and 3 (N=100).

In the full regression model, sex and education were the only significant predictors of SUS scores. Males rated usability an average of 7.3 points higher than females (95% CI: 0.4 to 14.2, p = 0.038). Participants with a graduate degree rated usability 13.3 points lower than those with less education (95% CI: −23.4 to −3.2, p = 0.010). SUS scores did not differ significantly by age, race, ethnicity, BMI category, U.S. region, survey round, or sampling protocol. Engagement scores were consistently high across groups, with no statistically significant differences by any demographic or study-related variable.

The mean rating for how helpful the onboarding tutorial was in providing an overview of the app was favorable, with a score of 3.9 (SD = 1.0) out of 5. Helpfulness of the video tutorials received a similarly favorable rating, with a mean of 3.8 (SD = 1.0) out of 5. The results were similar when analysed by round (all p ≥ 0.592).

### Qualitative Findings and *Edna* Modifications

**Table 3** presents the most frequently-cited usability issues reported by participants, along with illustrative feedback and the corresponding refinements made across development rounds.

**Table 3.** Top cited usability issues and refinement decisions.

| Usability Issue | Description | Example User Feedback | Refinement Made |
| --- | --- | --- | --- |
| **Limited food options** | Users requested more food variety, especially for cultural dishes, restaurant meals, and add-ons. | *"Include more diverse food options. I wanted to add a taco bowl or Korean BBQ."* | Reviewed missing foods and updated list; added aliases for difficult-to-find items (after rounds 1 and 2). |
| **Confusing food categorization** | Users struggled to locate foods in the list and didn't always understand how to browse categories. | *"Some foods were hard to find. For example, I expected to find burgers under fast food, not sandwiches."* | Introduced "Find My Food" feature with crumb trail (after round 1); reworded some category labels (after rounds 1 and 2). |
| **Desire for more guidance** | Participants wanted more support during food entry, especially at the beginning. | *"A guide to help during the first couple of days would be helpful."* | Created multiple new tutorial videos (e.g. complex meals, how to find foods) (after round 1). |
| **Bugs and crashes** | Users reported app freezing, crashing, or being slow to load. | *"App shuts down a lot." "Sometimes freezes, have to exit and re-open."* | Addressed crashes and bugs between rounds; improved app stability and speed (after rounds 1 and 2). |
| **Want to edit or add to events** | Users found it frustrating to start a new flow to add foods they forgot earlier. | *"I forgot to add a beverage to a meal but couldn't just add it."* | Enabled adding new foods to existing events without repeating the full entry process (after round 2). |
| **Calendar and feedback visibility** | Users wanted to know which days had been completed and see an overview of entries. | *"Would be nice to see what days I've logged."* | Updated calendar to show days with entries; added daily summary screen (after round 1). |
| **Time entry limitations** | Some users wanted to log the exact time they ate, rather than choosing from intervals. | *"Allow me to put in the actual time I ate."* | Switched from 15-minute intervals to clock-time entry (after round 2). |
| **Reminders and prompts** | Users missed some features because they didn't know where to find them or forgot they existed. | *"Didn't know I could favorite foods."* | Added 24-hour prompts after first use to highlight FAQs, videos, Favorites, and Find My Food (after round 2). |

Common challenges included confusion around food categorization, the absence of portion sizes, and difficulties locating specific foods. Many users also expressed a desire for more guidance, especially early in the logging process, and reported issues related to app stability, editing entries, and discovering key features.

Between rounds 1 and 2, *Edna* was refined to improve stability, clarity, and ease of use. Technical issues were addressed, and onboarding was updated to clearly explain the app’s purpose (i.e., focus on specific foods high in saturated fat or added sugar to support research). In parallel, tutorial videos and FAQs were expanded, and interface updates included improvements to previewing food items, a daily summary screen, and a visual calendar showing which days had entries. To reduce the number of difficult to find foods, *Edna* was improved in 3 ways: (1) food category labels were revised to highlight the location of foods, (2) the “Find My Food” feature was introduced as an in-app navigation assistance feature, and (3) generative AI was used to develop food aliases for cultural and regional foods (e.g., “pad Thai” = “Asian noodle dish”) that were embedded into the “Find My Food” feature.

Between rounds 2 and 3, changes focused on addressing the most persistent issues. Feature visibility was improved through contextual prompts after the first 24 hours of app use, and the food entry flow was revised to allow users to add items to existing meals without restarting the process. Additional, minor refinements included new food aliases and allowing users to enter exact clock times for meals.

Improvements made between rounds 1 and 2 resulted in the greatest observed improvements in usability with the highest SUS score in the final round of testing (**Table 2**). Similarly, the greatest reduction in the approximate number of difficult to find foods was observed between rounds 1 and 2 (Round 1: 100; Round 2: 62; Round 3: 65). The limited improvements in usability achieved between rounds 2 and 3, confirmed that additional rounds of refinements would no longer appreciably improve usability scores.

### Portion Size Estimation Task and *Edna* modifications

Of the 55 participants in Round 3, n=34/55 (61.8%) volunteered to provide feedback on the portion size selection demo in the app. Overall impressions were positive. Most participants, n=32/34 (94.1%), rated their overall experience using the app, including portion size selection, as *excellent* or *nearly excellent*. Satisfaction with the portion size selection process was also high, with n=22/34 (64.7%) reporting they were *very satisfied* or *satisfied*.

Portion size labels were generally clear: n=30/34 (88.2%) reported no foods with unclear or confusing labels. Selecting portion sizes required *very little* or *little* effort for n=26/34 (76.5%), and all participants, n=34/34 (100%), found the labels shown with portion size images (e.g., “small,” “medium,” “1 cup”) to be *understandable* or *very understandable*. Most participants (n=22/34, 64.7%) thought that the portion size options were well matched to the amounts of food they typically eat.

The only notable portion size entry error was made by 5 of the 9 participants randomized to the menu that included eggs (Day 1). After reconfiguring how eggs were displayed in the app, 4 additional participants who were subsequently randomized to Day 1 correctly entered the egg portion size into the *Edna* tool.

Qualitative feedback supported the positive quantitative findings. Most described the portion size selection process as “very easy to navigate” and “straightforward,” with several highlighting the usefulness of visual cues. One participant noted, “Seeing a visual was very helpful. Also having different options like ‘1 cup’ or ‘1 container’ made it feel easy.” Many reported no difficulties, while a few identified specific foods that were challenging to interpret. For example, one participant remarked that “cupcake portion pics included icing, but icing was available as an add-on”. Suggestions for improvement were minimal, and most participants provided no additional dislikes. Participant feedback informed the final design of *Edna’s* image-based portion size selection feature. The success of these design refinements was evident in that participants who had previously experienced common issues (e.g., entering the portion size for eggs) no longer encountered problems with the refined version.

## Discussion

This study evaluated the usability of *Edna*, an mEMDA platform developed to modernize population-level dietary surveillance. Across iterative rounds of national remote testing, *Edna* achieved above-average usability and strong participant engagement, demonstrating that a mobile, EMA-based tool can collect detailed dietary data feasibly and acceptably across diverse adults. Participants consistently rated the app as easy to use and intuitive, noting that the interface, visual cues, and structured logging flow supported accurate and efficient meal entry. Engagement remained high across the study, reflecting sustained motivation and minimal burden during daily use. These results demonstrate the feasibility of EMA-based, image-supported dietary surveillance in diverse U.S. adults and provide proof-of-concept for a scalable approach that integrates real-time self-report with automated nutrient coding.^24^

Results reinforced the feasibility of EMA-based dietary assessment. SUS scores rose from 71.7 to 78.8 in three rounds of iterative design and usability testing. These scores are well above the widely accepted “average” benchmark of 68 for digital tools^24,25^ and approached the range typical of well-designed health applications.^24^ Similarly, participants rated the image-assisted portion module highly for clarity and ease. These findings are consistent with prior evidence that image guidance improves portion estimation and reduces burden.^26^ The most substantial improvement to the SUS scores occurred between Rounds 1 and 2, coinciding with refinements to the leveled food-entry flow: simplifying food categorization, introducing a “Find My Food” helper with aliases and synonyms, enabling adding foods to existing meals, improving clock-time entry and calendar navigation, and surfacing Favorites for one-tap re-entry. These adjustments directly addressed early user feedback about navigation and edit burden, producing measurable gains in perceived usability. SUS scores plateaued after Round 2, suggesting that the interface had reached a stable and satisfactory level of usability where further gains would likely require micro-interaction or aesthetic refinements rather than structural redesign, despite falling short of the pre-defined A grade usability goal of 80.8 on the SUS.

Engagement, defined as the percentage of study days on which participants logged at least one food or beverage, was exceptionally high (≈ 90%) across rounds. Compliance, defined as the percentage of scheduled intervals completed within the interval-contingent group, was also strong where measured, ranging from 76% to 87% across rounds. Both metrics substantially exceed average EMA performance benchmarks, as meta-analytic findings place overall EMA compliance near 75% in substance-use studies^27^ and around 79% in general mobile-EMA designs.^28^ Importantly, usability, engagement, and compliance were comparable across event-and interval-contingent protocols, indicating that when user experience is well designed, either sampling scheme can sustain high participation. While this pattern needs to be replicated as a primary outcome, it does suggest that future studies might be able to base sampling-scheme selection on analytic priorities, such as capturing contextual richness versus estimating time-stamped prevalence, rather than on concerns about differential burden or dropout.

Strengths of the usability study include a priori usability benchmarks, randomized comparison of sampling protocols, national remote recruitment, and a multimethods approach linking concrete design refinements to measurable usability gains. Limitations should be considered including that participants were compensated for study involvement; however, two compensation schemes were used and did not meaningfully affect engagement. Compensation is standard and ethically expected in EMA research, and our approach was consistent with prior practice.^29^ The study was conducted in English only and future design should consider multi-language applications. However, *Edna’s* inclusive food list, visual design, and integration of cultural food aliases into the “Find My Food” feature minimized literacy demands, partially mitigating linguistic constraints. Future versions will offer multilingual interfaces. Usability reflects perceived experience rather than objective accuracy; however, a structured portion-size simulation was incorporated to assess navigation and comprehension of image cues, providing early evidence of functional validity. Further nutrient coverage was limited to saturated fat and added sugars; however, this focused scope enabled rigorous usability testing before expanding to broader nutrient domains.

## Conclusions

Collectively, these findings demonstrate that mobile ecological momentary diet assessment is both feasible and acceptable for population-level dietary surveillance when grounded in iterative, user-centered design. By combining real-time self-report, visual portion guidance, and automated nutrient coding within an accessible interface, *Edna* provides a practical bridge between traditional 24-hour recalls and emerging digital surveillance methods. As public health agencies seek scalable approaches that capture diet more dynamically and inclusively, *Edna* offers a foundation for integrating mEMDA into national nutrition monitoring systems to strengthen evidence-based efforts to improve diet quality and reduce nutrition-related inequities.

## Future Directions

Overall usability was strong; however, a few participants indicated that the leveled food-entry flow could be further streamlined. To modernize *Edna’s* interface and further enhance efficiency, future development efforts will integrate AI-assisted search and adaptive prompting to make food identification faster and more intuitive. The next phase will also validate *Edna* against known foods to quantify accuracy in food identification and portion selection, and conduct a free-living comparison of event- versus interval-contingent sampling to identify which approach yields the strongest compliance, engagement, and recall fidelity in naturalistic conditions. These efforts will extend *Edna’s* capability as a scalable, user-centered platform for modern dietary surveillance.

## Author contributions

SMS conceptualized and supervised the study and obtained funding. MJ conducted the statistical analysis and prepared the visualizations. RW produced software to collect the data. EJB provided methodological support. JS, and JF contributed to data collection. KMR, CAT, GD, CT contributed to study conceptualization. SMS and MRJ wrote the initial manuscript draft and all authors read and approved the final manuscript.

## Data Availability

The datasets generated and analysed during the current study are available in the Zenodo repository, 10.5281/zenodo.17538030.

## Acknowledgements

This research was funded by National Institutes of Health, National Cancer Institution, grant number R01CA244404. The funder played no role in study design, data collection, analysis and interpretation of data, or the writing of this manuscript.

## Competing interests

MRJ declares current consultation to WW International, Inc. and past consultation to ZOE. KMR declares current employment at WW International, Inc. RW declares employment by Viocare Inc. All other authors declare no competing interests.

## Notes

### Author Declarations

Institutional Review Board of Georgetown University gave ethical approval for this work.

